# Evaluation of Physico-chemical parameters of selected Clotrimazole pessaries from pharmaceutical importers and Accredited Drug Dispensing Outlets in Dar es Salaam, Tanzania

**DOI:** 10.1101/2025.01.21.25320887

**Authors:** Benedict Brashi, Eulambius M. Mlugu, Eliangiringa A. Kaale

## Abstract

**Introduction:** Quality assured medicines make an important contribution to a global reduction in morbidity and mortality. Tanzania depends on approximately 70% of imported medicines, some are manufactured in countries with different climatic zones for stabilities. The prevalence of vaginal candidiasis amongst women seeking primary health care for genital infections in Dar es Salaam is 45%. Clotrimazole pessaries is the first-line drug for the treatment vaginal candidiasis (VC). However, their reports show the existence of poor quality clotrimazole in the market as well as microbial resistance.

**Study objectives:** This study aimed to assess the quality of clotrimazole pessaries among pharmaceutical importers and Accredited Drug Dispensing Outlets (ADDOs) outlets in Dar es Salaam.

**Methods:** The experimental study laboratory-based was conducted between October 2021 to July 2022. A total of 72 samples, 5 from pharmaceutical importers and 67 from ADDO outlets, were collected by mystery clients through the convenient method. Assay for the content of active ingredients and disintegration time were performed as described in the British Pharmacopoeia (BP).

**Results:** All samples passed the physical inspection and disintegration test. Among the collected samples, two of 72(2.78%) failed the assay test. The assay for an active pharmaceutical ingredient for samples from importers was significantly greater than that from ADDO outlets.

**Conclusion:** Storage conditions may affect the drug content of clotrimazole pessaries. A few samples from ADDO without fan failed to meet the assay test as per British Pharmacopeia specification.

**Recommendations:** The Pharmacy Regulation 2019, for Accredited Drugs, Dispensing Outlets, Standards, and Ethics for Dispensation of Medicines to be revitalized to include a specification on ADDO outlets regarding the storage conditions to align with the pharmaceutical manufacturer’s recommendations. Also, regulatory inspections should be emphasized by the Regulatory Authority including Pharmacy Council and TMDA so as ensure the registered pharmaceutical outlets continue complying with standards required like the presence of air conditions so that the medicines continue to be of desired quality throughout their shelf life.

## 1. Introduction

Vaginal candidiasis is among the diseases affecting women, especially in resource-limited countries. For example, in Dar es Salaam Tanzania, 45% of women seeking primary care for genital infections had vaginal candidiasis (VC)(1). Even though these infections do not result in high mortality, but can be long-lasting and difficult to cure; this can result in high levels of anxiety and decrease the quality of life(2,3). Clotrimazole is among the drugs of choice to treat vaginal candidiasis (4,5). Clotrimazole belongs to Biopharmaceutical Classification System (BCS) class II, exhibiting low solubility. Its chemical structure is indicated in **Figure I**.

**Figure 1.**
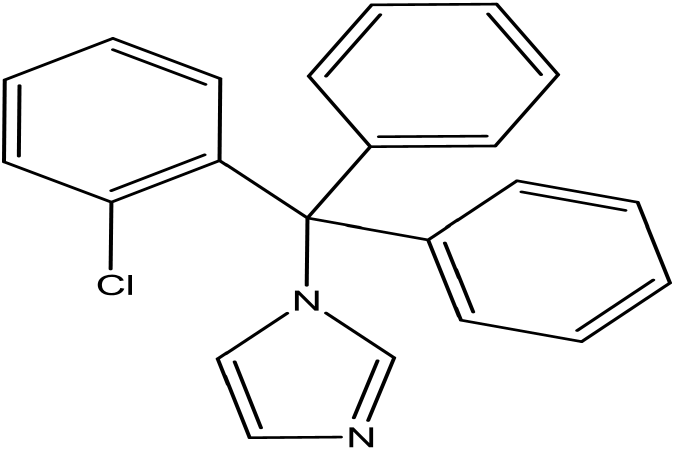
Chemical Formulae of Clotrimazole (C_22_H_17_ClN_2_)

Vaginal drug delivery presents several advantages, including ease of administration, hepatic first pass-effect, low systemic drug exposure, and increased permeability for some drugs compared to oral or other routes(6,7).

The quality of medicines imported into tropical climate countries may be adversely affected if their formulations are not being optimized for stability under Zone four B (IV B) conditions. The affected parameters include active ingredient content and disintegration time(6). Thus, there is a concern in verifying the quality of medicines at the end-user level.

The study in the United States, conducted on medicinal products stored at high temperature and humidity, demonstrated a substantial increase in formulation’s disintegration time, which is accounted for by moisture leading to hardening of the formulation. Thus, due to a rise in disintegration time, the formulation may fail to release the medicament resulting in treatment failure (8)

The study reported polymorphic and crystal formation in New Zealand, which involved formulations exposed to 23 °C, 30 °C and 4,0 °C with relative humidity of 75%RH and 96%RH for one month. The study explained that when the medicinal products are stored on premises with poorly controlled conditions like humidity and temperature, the medicaments can undergo polymorphic or crystal changes that may affect their inherent characteristics like solubility and thus can have increased time of disintegration (9)

In Saudi Arabia, a study that investigated the effect of quality of medicines on storage conditions reported that pharmaceutical products with excipients like lactose when stored in premises with high humidity and temperature, increase hardness and disintegration time; the probable reason is increased interparticle bonding by partial surface solution and recrystallization of the base and thus may affect the therapeutic effect (10)

A study in Nigeria reported a failure rate of 21.05% for the disintegration time of pharmaceutical products marketed in Edo, Anambra, and Delta states. One of the explanation on the failure was poor storage conditions like storing the medicines under high relative humidity or high temperature which may interfere with the properties of the binders and disintegrants thus affecting the medicament release (11)

In Tanzania, Accredited Drug Dispensing Outlets (ADDO) program was established in 2003, to improve access to essential medicines in peri-urban and rural areas with no pharmacies. However, there are reports on poor storage facilities identified as among the challenges facing private ADDO(12). This in turn might lower the quality of medicines available in these shops.

The study in Tanzania reported 12.2% failure in the quality of antimalarial in retail pharmacies (13) and another study in Nigerian pharmacies revealed 48% non-compliance to pharmacopeia specifications (14). This divulgence recapitulates the importance of monitoring pharmaceutical product quality to safeguard public health.

Clotrimazole pessaries are among the medicines which have shown poor quality and safety concerns in the Tanzania pharmaceutical market (15). The reported quality problems include failure of the Clotrimazole pessaries to dissolve when inserted in the vaginal. Also, some studies in Kenya have reported the resistance of microbial isolates to Clotrimazole pessaries (16–18). Globally, various studies to evaluate the quality of medicines have been undertaken and data on quality is available. But due to a limited number of research in assessing the quality of intravaginal formulations in tropical conditions, thus there is little data on their performance in the tropical climatic market (6). Also, there is no data on clotrimazole pessaries sold in ADDO outlets in Dar es Salaam.

Therefore, the purpose of this study was to assess the quality of clotrimazole pessaries. Also, to investigate the influence of storage conditions on the physicochemical parameters of Clotrimazole pessaries in the Dar es Salaam market.

## 2. Materials and Methods

### 2.1. Study site

This experimental study laboratory-based was conducted in 2022 in the Dar es Salaam region. The region was selected due to its tropical climatic conditions (hot and humid throughout the year), highly populated, and the prevalence of VC is as high as 45% (3). Also, the city is a business hub where the five Clotrimazole pessaries importers are located(19). The samples were analyzed by the TMDA Quality Control Laboratory, a WHO prequalified for chemical testing of medicines.

### 2.2. Sample collection

The study involved all five registered brands of clotrimazole pessaries present in the Tanzania market, including labesten, clotrine, clorifort, candid V6, and candistat pessaries. A total of 72 samples from five brands of clotrimazole pessaries were available in the study area. The sampling used was convenient where all the batches present in the market with a minimum of 26 pessaries were collected. The sampling sites considered the proportion and distance of ADDO from town to outside the city as the population varies from City Centre to area outside the city.

### 2.3. Chemical and Reagents

Methanol (HPLC grade) from MRS Scientific Ltd (Unit 5-Wickford, United Kingdom), ortho-phosphoric acid (HPLC grade) from Merck KGaA (Darmstadt, Germany), triethylamine (HPLC grade) from Scharlab S.L (Gato Perez, Spain). Clotrimazole CRS was procured from the European Directorate for the Quality of Medicines (EDQM) and HealthCare (Strasbourg, France).

### 2.4. Instrumentation

The disintegration test of tablets was conducted on DT ZT 53 disintegration tester from Erweka GmbH (Hausenstamm, Germany) while the assay test was conducted by using HPLC systems which were equipped with an online degasser, a binary pump, an automatic sample injector, a column oven, and a photodiode array detection module (all Shimadzu, Tokyo, Japan). The study involved the use of other equipment including weights of tablets and reagents were measured using AUW 120 balance from Shimadzu Corporation (Kyoto, Japan). Mettler Toledo pH and conductivity meter, an Ultra sonicator Branson 5800 from Branson Ultrasonics corporation (Danbury, United States of America). The samples including reference standards were filtered by using 0.45µm syringe filters before injection into the HPLC system. The study utilized type A glassware. Purified water for mobile phase and buffer preparation was generated by a Zero B Eco smart Reverse Osmosis from Ion Exchange LTD (India) and further purified by Ultra clear™ 10TWF 30 UV UF TM from Evoqua water Technology (Bursbuttel, Germany). Disintegration time was done by DT Erweka GmbH, Germany, and assay tests were done by HPLC employing a C-18 column with integrated precolumn (4.6 × 200mm, C18, 5µm particle size spherisorb ODS 1) (Waters, Ireland), is a silica-based, RP and non-end-capped packed with octadecylsilyl silica gel for the stationary phase.

### 2.5. Preparation of the mobile phase

The mobile phase preparation involved measuring and mixing 700mL of methanol and 300mL of 0.02M of ortho-phosphoric acid into 1000 mL Volumetric Flask (VF). pH was adjusted to 7.5 ±0.1 by using a 10% v/v solution of triethylamine in methanol. After pH adjustment, the mobile phase was then degassed for 20 minutes by using an ultra sonicator.

### 2.6. Preparation of buffer solution for adjusting pH of mobile phase

Solution of 10% v/v of solution of triethylamine in methanol was prepared by accurately measuring 10mL of triethylamine and 90mL of methanol.

### 2.7. System suitability solution

A concentration of 0.04mg/mL was prepared to determine the column efficiency by accurately weighing and dissolving 20mg of the clotrimazole reference standard in 100mL VF with methanol and water to a ratio of 70:30. Then, 1mL of the resulting solution was transferred into 5mL VF, and diluted to a mark by a mixture of methanol and water to a ratio of 70:30. The principal chromatogram obtained with this solution should be not less than (NLT) 9,000 theoretical plates per meter.

### 2.3 Preparation of sample solution for the content determination

Twenty pessaries of the same batch for each sample were weighed and powdered using mortar and pestle, then weight equivalent to 20mg was weighed using the weighing boat and placed into a 50mL VF, approximately 25mL of methanol was added and sonicated for 20 minutes. Then methanol was added to fill to mark of the VF, sonicated again for 5 minutes, and allowed to cool. The solution was then filtered into a conical flask, and then 5mL was pipetted into a 50mL VF. To VF 50mL of methanol and water to a ratio of 60:40 to make up a volume of 50mL. The resulting solution was shaken and mixed to ensure no lumps were present, then filtered by using a 0.45-micron filter and transferred into an HPLC vial where an injection of 20 µL was made.

### 2.4 Preparation of standard solution for content determination

Preparation of Clotrimazole reference standard was done by accurately weighing 5.0mg using a weighing boat and transferring into 25 mL VF. Next, added approximately 15 mL of a mixture of methanol and water to a ratio of 70:30, sonicated for 20 minutes diluted to volume with the same solvent. Next, 1mL of the resulting solution was pipetted into 5 mL VF and diluted to volume with the same solvent. Then filtered by using a 0.45-micron filter and transferred into an HPLC vial where an injection of 20 µL was made.

### 2.5 Disintegration test

The disintegration time of pessaries was determined using the BP method. The test was conducted on DT ZT 53 disintegration tester from Erweka GmbH (Hausenstamm, Germany). Initially purified water was filled into the tank to mark, then accurately measured 900mL of purified water into a beaker as disintegration media into the two (2) disintegration vessels, and the circulator was switched on. The medium was heated to 37^0^C; the external thermometer was used to verify the media temperature was maintained at 37±0.5^°^C. Six pessaries from each batch were individually transferred into a disintegration vessel; the time was set at 30 minutes as specified in the monograph and operated. The observation of pessaries in DT apparatus after every 10 seconds and recorded when completely disintegrated.

### 2.6 Method Verification

The Clotrimazole pessaries BP monograph was verified to fulfill the requirements of GMP and pharmacopeial implementation to ascertain the applicability of method transfer. Without a placebo, the method was verified for system precision, linearity, and accuracy parameters. Then, the method was used to determine the content of clotrimazole API from the clotrimazole pessaries with reference limits of 100±5% (20).

## 3. Method Verification

The Clotrimazole pessaries BP monograph was verified to fulfill the requirements of GMP and pharmacopeial implementation to ascertain the applicability of method transfer. Without a placebo, the method was verified for system precision, linearity, and accuracy parameters. Then, the method was used to determine the content of clotrimazole API from the clotrimazole pessaries with reference limits of 100±5% (20).

### 3.1. Linearity

To achieve the calibration curves, the calibration solutions were prepared by accurately weighing 5mg of Clotrimazole using the weighing boat and transferring into 25 mL VF. Approximately 15ml of methanol and water to a ratio of 70:30 was added and sonicated. After sonication, the same solvent was added to the mark of VF, sonicated again for 5 minutes, then the solution was filtered and a correct volume of the solution was accurately measured and diluted accordingly to achieve the concentration of 50%, 80%, 100%, 120%, and 150% of five (5) calibration solutions. The resulting solution was filtered, transferred into HPLC vials, and then injected in duplicate. The linearity was evaluated visually from plotted calibration curve as shown in **Figure 2**.

**Figure 2.**
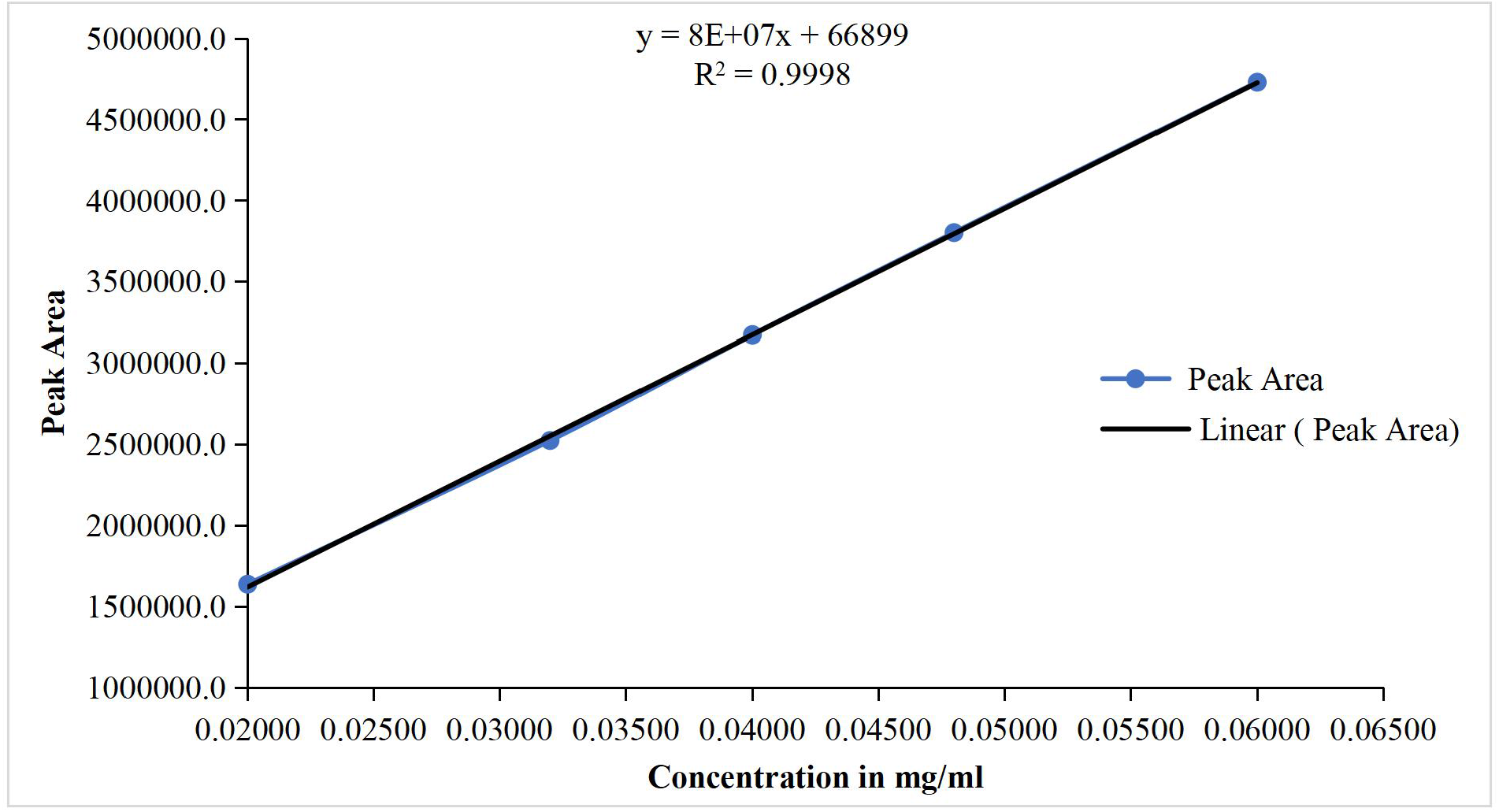
Calibration curve indicating a linear relationship between concentration and peak area response for Clotrimazole

### 3.2. System precision

Precision in terms of repeatability was investigated by independently preparing 5 calibration solutions. The solution was prepared by accurately weighing 5mg of Clotrimazole using weighing boat and transferred into 25 mL VF and approximately 15ml of methanol and water to a ratio of 70:30 was added and sonicated. After sonication, the same solvent was added to the mark of VF, sonicated again for 5 minutes, then the solution was filtered and a correct volume of the solution was accurately measured and diluted accordingly to achieve the concentration of 50%, 80%, 100%, 120%, and 150% of 5 calibration solutions. The resulting solution was filtered, transferred into HPLC vials, and then injected in duplicate. Regression statistics and regression analysis indicated the fitness of the regression model establishing a linear relationship between concentration and peak response as shown in **Table 1**, then, standard deviation, relative standard deviation, and recovery were determined.

**Table 1.** Assay test results for accuracy without placebo.

| Sample | Percentage Recovery (98%-102%) | Average | Standard Deviation | %RSD |
| --- | --- | --- | --- | --- |
| 80% (S1) | 99.6 | 100.1 | 0.456 | 0.5 |
| 80% (S2) | 100.2 |  |  |  |
| 80% (S3) | 100.5 |  |  |  |
| 100% (S1) | 99.6 | 100.1 | 0.456 | 0.5 |
| 100% (S2) | 100.2 |  |  |  |
| 100% (S3) | 100.5 |  |  |  |
| 120% (S1) | 102.0 | 102.0 | 0.098 | 0.1 |
| 120% (S2) | 102.0 |  |  |  |
| 120% (S3) | 102.1 |  |  |  |

### 3.3. Accuracy

The calibration solutions were prepared by accurate weighing the respective amount of reference standard and mixing with the required number of clotrimazole pessaries tablets in 50 mL VF, diluted and sonicated, and filled the VF to mark. The accuracy was investigated at 80%, 100%, and 120% of the calibration levels described above under 2.4 and evaluated the recovery of the analyte at each level. The percentage of matrix used in accuracy was 106.8% and the purity of CRS was 99.8mg/g. The results were summarized in **Table 1** above.

### 3.4. Data management and analysis

The data from the field and the lab testing were entered, cleaned, and analyzed using Statistical Package for the Social Science (SPSS) Version 26. Numeric variables were summarized using mean and standard deviation. Frequency and proportions were summarized as categorical variables. The comparison among sample sources was conducted by One-way Analysis of Variance (ANOVA) was performed to evaluate whether exists a significant difference in disintegration tests and assay test results within suppliers and among suppliers, p-value <0.05 was considered a significant level

## 4. Results

All samples passed the physical inspection and disintegration test. Among the collected samples, two of 72(2.78%) failed to meet the specifications of the assay test. The assay for an active pharmaceutical ingredient for samples from importers was significantly greater than that from ADDO outlets.

### 4.1. Method verification

#### 4.1.1. Linearity

The method was linear in the concentration range of 0.120 – 0.380 mg/mL. The linear regression of 0.9998 was obtained. The linearity was evaluated visually from plotted calibration curve as shown in **Figure 2**. This indicated that there is a linear relationship between the concentration and response in terms of peak area.

#### 4.1.2. Precision

In the determination of system precision, repeatability for the three batches investigated ranged from 0.1– 0.5 percentage relative standard deviation (%RSD) which was within the acceptable limit of NMT 2.0%. Similarly, recovery ranged from 100.1 - 102%, which was within the acceptable limit of 98.0 – 102.0% **Table 1**.

#### 4.1.3. Accuracy

The overall recovery results for the investigated levels which were 80%, 100%, and 120%, ranged from 98.0 – 102.0%, which was within the acceptable limit of 98.0 – 102.0%.

#### 4.1.4. Analysis of Clotrimazole pessaries samples

All 72 samples of clotrimazole pessaries were subjected to quality control testing. The investigated parameters include visual/physical inspection, disintegration time, and assay against the requirements prescribed under British Pharmacopoeia(20).

##### 4.1.4.1. System Suitability test

The theoretical plates per meter of 18,674.091 was attained while the specification was NLT 9,000 theoretical plates per meter as BP. These results indicate that all requirements of the system suitability solution were achieved and thus allowing the content determination to commence(20).

##### 4.1.4.2. Appearance/visual inspections

All 72 clotrimazole pessaries samples complied with packaging and labeling requirements. The labels of the collected clotrimazole samples contained instructions to store below 30^°^C, protect the product from light, and to avoid freezing. The manufacturers’ instructions on storage temperature were consistent with all samples from all five manufacturers.

##### 4.1.4.3. The storage conditions at sampling sites

During sample collection, the premises temperature and humidity were recorded as prescribed in sample collection forms. An overview of the recorded storage conditions in the sampling sites is presented in **Table 2**. The compliance with the storage conditions declared by manufacturers was generally good for importers and for ADDO the temperature of 31^°^C was recorded, slightly above the recommended storage conditions for products by manufacturers.

**Table 2.**
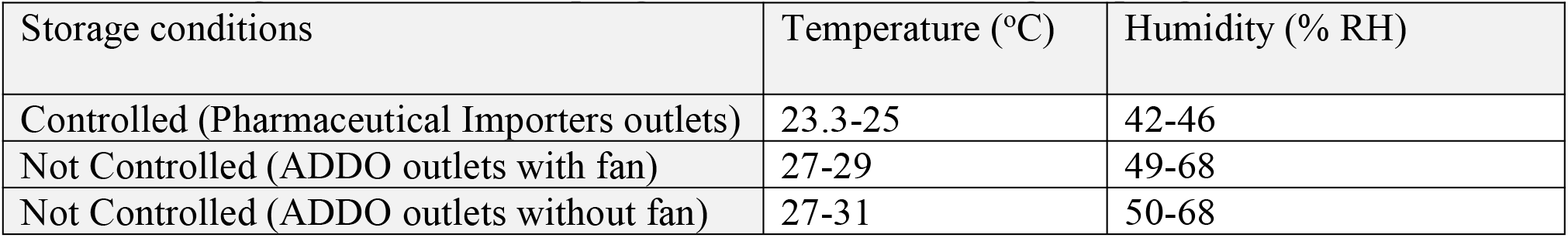
Storage conditions at sampling sites, as recorded during sampling.

##### 4.1.4.4. Disintegration time

The mean disintegration time with one SD for clotrimazole pessaries from pharmaceutical importers, ADDO outlets with fan, and ADDO outlets without a fan was 2.75(1.17), 5.42(4.23), and 7.95(6.28) minutes respectively. All the samples complied with requirements prescribed under BP 2020 as summarized in **Table 3**.

**Table 3.**
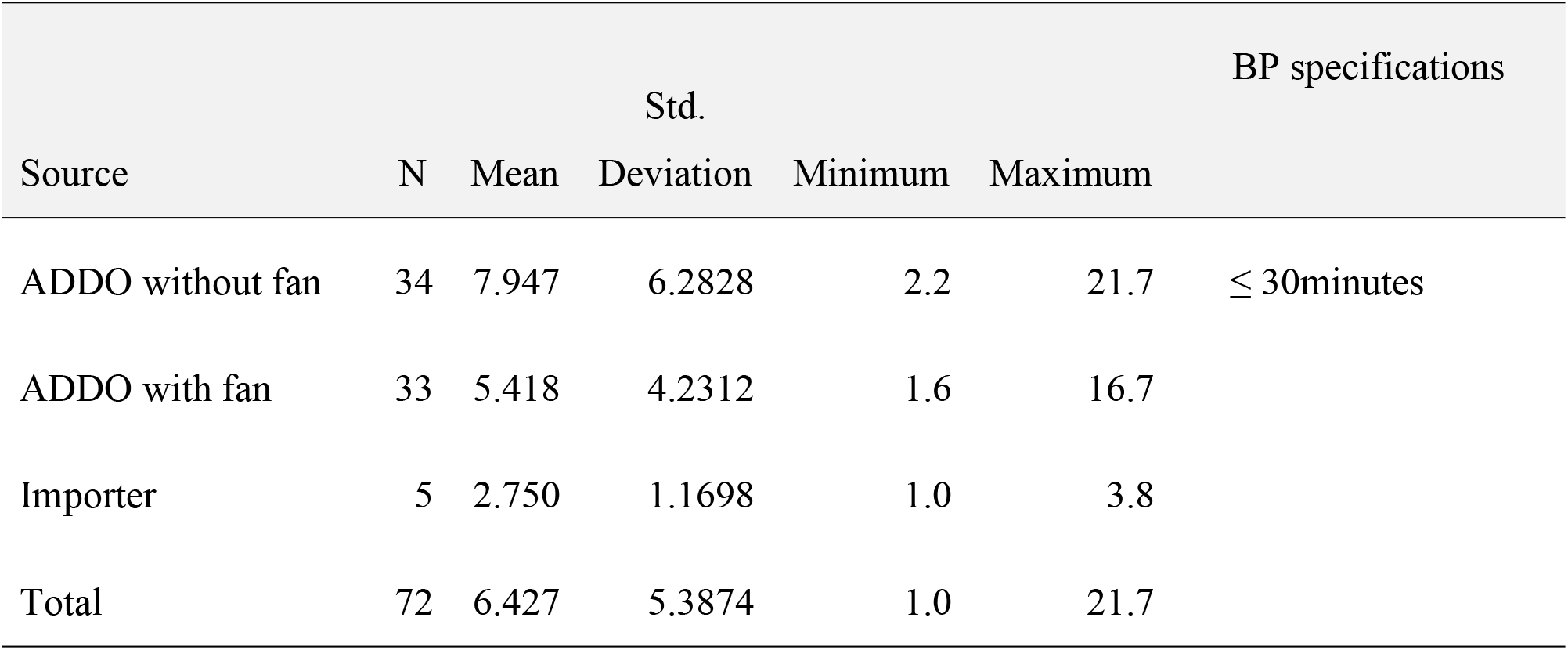
Disintegration test results with respect to pharmacopeia specification (n=72)

##### 4.1.4.5. Assay

The mean API content for 20 tablets with one SD for clotrimazole pessaries from pharmaceutical importers, ADDO outlets with fan, and ADDO outlets without a fan was 102.46% (1.49%), 98.85% (1.77%) and 97.47(1.890%) respectively. On average all the samples passed the assay test except 2 of 72(2.78%) from ADDO outlets without fans failed to comply with BP specifications as detailed in **Table 4**.

**Table 4.** Assay test results with respect to pharmacopeia specifications from three sources(n=72)

| Source of a drug | N | Mean | Std. | Minimum | Maximum | BP |
| --- | --- | --- | --- | --- | --- | --- |
|  |  | (%) | Deviation |  |  | specification |
| ADDO without fan | 34 | 97.471 | 1.8891 | 94.2 | 100.3 |  |
| ADDO with fan | 33 | 98.848 | 1.7699 | 95.1 | 101.9 | 95-105% |
| Importer | 5 | 102.460 | 1.4943 | 101.2 | 105.0 |  |
| Total | 72 | 98.449 | 2.2050 | 94.2 | 105.0 |  |

On evaluation, whether the means from all three sample sources were statistically different One-way ANOVA was used. From this test, it was found that the assay of API at importers was 3.61% greater than that of ADDO with fans and was statistically significant (p=0.001). Also, the assay content at the importer was 4.99% significantly greater than that at ADDO outlets without a fan (p=0.001). Moreover, the assay content of API at ADDO outlets with the fans was found to be 1.38% greater than that at ADDO outlets without a fan, and it was statistically significant (p=0.008). The details as shown in **Table 5**.

**Table 5.** Differences in mean assay percentage between a sample from three sources.

| (I) Source | (J) Comparison category | Mean Difference (I-J) | Std. Error | Sig. |
| --- | --- | --- | --- | --- |
| ADDO without fan | ADDO with fan | -1.3779* | 0.4432 | 0.008 |
|  | Importer | -4.9894* | 0.8686 | 0.000 |
| ADDO with fan | Importer | -3.6115* | 0.8703 | 0.000 |

## 5. Discussion

In this study, it was found that disintegration time for Clotrimazole pessaries of different brands collected from ADDO outlets with fans and without fans was not significantly different from that of samples collected from pharmaceutical importers, and all complied with requirements prescribed under BP 2020(20).

The conformity of the Clotrimazole pessaries to the British Pharmacopeia specification for disintegration time can be explained as the result of the appropriately use of disintegrants and excipients like binders by the manufacturing companies(11)

The findings from this study were similar to those found in the study in Northern Myanmar(21). Also, a study in Nepal examined the physicochemical parameters of multinational medicines in the market, where all the samples passed the disintegration test (22).

The current study revealed that 2 of 72(2.78%) collected samples failed the assay test as per specifications of BP (95-105%. The samples were collected from ADDO outlets without a fan. The explanation for the low assay content may be due to poor storage conditions. The assay of API from importers was 3.61% significantly greater than that of ADDO with fan(p=0.001). Also, the assay content at the importer was 4.99% significantly greater than that at ADDO outlets without a fan (p=0.001). Moreover, the assay content of API at ADDO outlets with the fan was found to be 1.38% greater than that at ADDO outlets without a fan, and it was statistically significant (p=0.008).

The findings of our study were near agreement to the study conducted in Tanzania, which involved samples collected from ADDOs and pharmacies for other medicines, which revealed a failure rate for assay tests to be two samples with 6.6% failed assay (23). In the cited study, the reason for the failure was stated to be the stability of formulation to tropical temperatures, and light, which amounts to the lack of adherence to good storage practices was pointed out as a reason.

For antimicrobial drug-like clotrimazole to work effectively, it must contain the active pharmaceutical ingredient in the recommended content, that is 95-105% of the label claim, the use of antimicrobial with a low amount of API can lead to antimicrobial resistance. Several studies in Kenya have published data on the resistance of microbes to clotrimazole pessaries (16– 18). With other steps being taken to combat antimicrobial resistance, the enforcement to ensure premises where the medicines are stored need to be regularly enforced to ensure the medication retains its quality throughout its shelf life.

## 6. Conclusion

Storage conditions may affect the drug content of clotrimazole pessaries. Among the samples collected 2.78% from ADDO without fan failed to meet the assay test as per British Pharmacopeia specification.

## Data Availability

All data produced in the present study are available upon reasonable request to the authors

## 7. Recommendations

The Pharmacy Regulation 2019, for Accredited Drugs, Dispensing Outlets, Standards, and Ethics for Dispensation of Medicines to be revitalized to include a specification on ADDO outlets regarding the storage conditions to align with the pharmaceutical manufacturer’s recommendations. Also, regulatory inspections should be emphasized by the Regulatory Authority including Pharmacy Council and TMDA so as ensure the registered pharmaceutical outlets continue complying with standards required like the presence of air conditions so that the medicines continue to be of desired quality throughout their shelf life.

## 8. Ethical consideration

The study was conducted after being granted approval number MUHAS-REC-04-2022-1081 from the Muhimbili University of Health and Allied Sciences Research and Publication Committee. This research was conducted following MUHAS ethical regulations and requirements.

## 10. Acknowledgments

The authors are indebted to Mr. Fabian Boniface from the Department of Medicines and Complementary product analysis for the technical and intellectual support when conducting this work.

